# Global profiling of SARS-CoV-2 specific IgG/ IgM responses of convalescents using a proteome microarray

**DOI:** 10.1101/2020.03.20.20039495

**Authors:** He-wei Jiang, Yang Li, Hai-nan Zhang, Wei Wang, Dong Men, Xiao Yang, Huan Qi, Jie Zhou, Sheng-ce Tao

## Abstract

COVID-19 is caused by SARS-CoV-2, and has become a global pandemic. There is no highly effective medicine or vaccine, most of the patients were recovered by their own immune response, especially the virus specific IgG and IgM responses. However, the IgG/ IgM responses is barely known. To enable the global understanding of SARS-CoV-2 specific IgG/ IgM responses, a SARS-CoV-2 proteome microarray with 18 out of the 28 predicted proteins was constructed. The microarray was applied to profile the IgG/ IgM responses with 29 convalescent sera. The results suggest that at the convalescent phase 100% of patients had IgG/ IgM responses to SARS-CoV-2, especially to protein N, S1 but not S2. S1 purified from mammalian cell demonstrated the highest performance to differentiate COVID-19 patients from controls. Besides protein N and S1, significant antibody responses to ORF9b and NSP5 were also identified. In-depth analysis showed that the level of S1 IgG positively correlate to age and the level of LDH (lactate dehydrogenase), especially for women, while the level of S1 IgG negatively correlate to Ly% (Lymphocyte percentage). This study presents the first whole picture of the SARS-CoV-2 specific IgG/ IgM responses, and provides insights to develop precise immuno-diagnostics, effective treatment and vaccine.

**Highlights:**

- A SARS-CoV-2 proteome microarray contains 18 of the 28 predicted proteins
- The 1^st^ global picture of the SARS-CoV-2 specific IgG/ IgM response reveals that at the convalescent phase, 100% of patients have IgG/ IgM responses to protein N and S1
- Significant antibody responses against ORF9b and NSP5 were identified
- Protein S1 specific IgG positively correlates to age and LDH, while negatively to Ly%

## Introduction

COVID-19 is caused by coronavirus SARS-CoV-2^1,2^. In China alone, by Mar. 18, 2020, there are 81,163 diagnosed cases with SARS-CoV-2 infection, and 3,242 death according to Chinese CDC (http://2019ncov.chinacdc.cn/2019-nCoV/). Globally, 184,976 diagnosed cases were reported in 159 countries, areas or territories. And on Mar. 11, WHO announced COVID-19 as a global pandemic. Aided by the high-throughput power of Next Generation Sequencing (NGS), the causative agent of COVID, *i*.*e*., SARS-CoV-2, was successfully identified and genome sequenced. Sequence analysis suggests that SARS-CoV-2 is most closely related to BatCoV RaTG13 and belongs to subgenus Sarbecovirus of Betacoronavirus, together with Bat-SARS-like coronavirus and SARS coronavirus^1,2^. Through the comparison with SARS-CoV and other related coronaviruses, it is predicted that there are 28 proteins may encoded by the genome of SARS-CoV-2, including 5 structure proteins (we split S protein to S1 and S2, and thus count as 2 proteins), 8 accessory proteins and 15 non-structural proteins^3^. SARS-CoV-2 may utilize the same mechanism to enter the host cells, *i*.*e*., the high affinity binding between the receptor binding domain (RBD) of the spike protein and angiotensin converting enzyme 2 (ACE2)^4-9^.

Though tremendous efforts are being poured for hunting effective therapeutic agents, *i*.*e*., small molecules/ neutralization antibodies, and protective vaccines, unfortunately, none of them are available at this moment, even in the near future^10^. By Mar. 18, 2020, 69,740 patients have been cured in China (http://2019ncov.chinacdc.cn/2019-nCoV/). Since there is no effective anti-SARS-CoV-2 drug and therapeutic antibody, theoretically, most of these patients are cured by themselves, *i*.*e*., by their own immune system. It is known that for combating virus infections, usually antibodies (IgG and IgM) play critical roles, for example, SARS-CoV^11,12^ and MERS-CoV^13,14^. Thus, it is reasonable to argue that virus specific IgG and IgM may also significantly contribute to the battle against SARS-CoV-2 infection. Indeed, high levels of SARS-CoV-2 specific IgG and IgM could be monitored for many of the patients^15^. In addition, positive results were observed by treating the patients with convalescent plasma collected from COVID-19 patients^16,17^.

However, because SARS-CoV-2 is a newly occurred pathogen, the IgG/IgM response is barely known. There are many important questions need to be experimentally addressed: 1. What’s the variation among different patients, especially for antibodies against nucleocapsid protein (protein N) and spike protein (protein S)? 2. Besides nucleocapsid protein and spike protein, is there any other viral protein that could trigger significant antibody response for at least some of the patients? 3. Is it possible to link the intensity of the overall IgG/IgM response to the severity of patients? and etc. It is urgently needed to answer these questions, especially in a systematic manner. Once these questions are answered or at least touched, we may can understand the IgG/IgM response in detail, and in turn facilitate us to develop more effective treatment, therapeutic antibody and protective vaccine.

Traditional techniques for studying IgG/ IgM responses including ELISA^18-20^, and immune-colloidal gold strip assay^19,21,22^. However, these techniques usually can only test one target protein or one antibody in one reaction. Thus a powerful tool is needed that enables the studying of the IgG/ IgM responses on a systems level. Featured by the capability of high-throughput and parallel analysis, and miniaturized size, protein microarray may be the choice for systematic study of the SARS-CoV-2 stimulated IgG/IgM responses. A variety of protein microarrays have already been constructed and successfully applied for serum antibody profiling, such as the Mtb proteome microarray^23^, the SARS-CoV protein microarray^11^, and the Dengue virus protein microarray^24^.

Here, we present a SARS-CoV-2 proteome microarray developed using an *E*.*coli* expression system and SARS-CoV-2 proteins collected from several commercial sources. COVID-19 Convalescent sera were analyzed on the microarray, the first overall picture of SARS-CoV-2 specific IgG/ IgM responses was revealed.

## Results

### Schematic diagram and workflow

The genome of SARS-CoV-2 is ∼29.8 kb, which is predicted to encode 28 proteins^3^, *i*.*e*., 4 structural proteins (split S protein as S1 and S2), 8 accessory proteins and 15 non-structural proteins (nsp) (**Fig. 1A**). The sequences of all these proteins and the receptor binding domain (RBD) on S1 were codon optimized, gene synthesized and cloned to appropriate vector for expression in *E. coli*. We affinity purified these proteins, meanwhile, we collected recombinant proteins expressed from both prokaryotic and eukaryotic systems from other sources. After quality control, these proteins were then printed on appropriate substrate slides. Convalescent sera were collected and analyzed on the proteome microarray. The global SARS-CoV-2 specific IgG and IgM responses were revealed.

**Figure 1.**
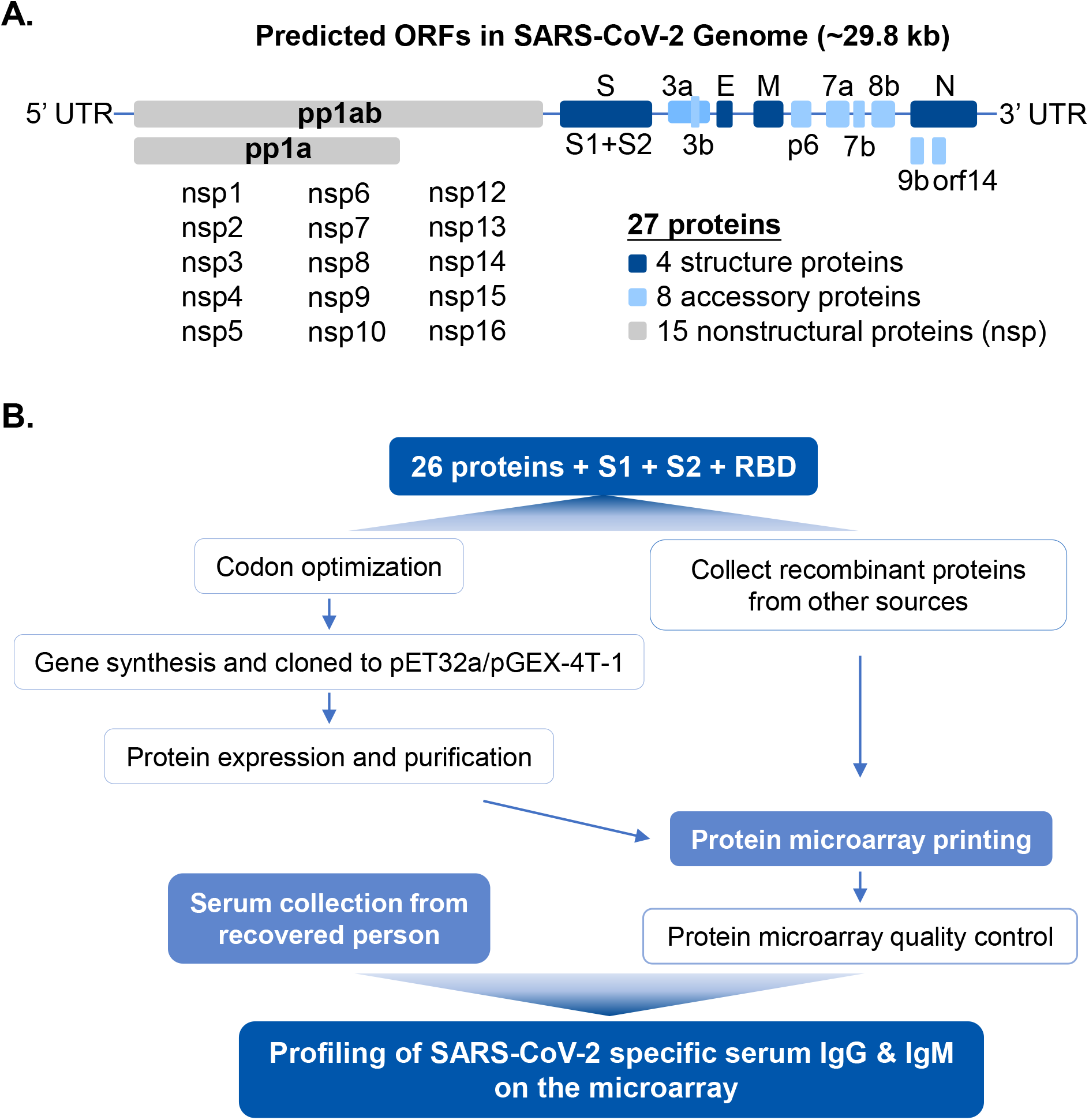
The workflow of SARS-CoV-2 proteome microarray fabrication and serum profiling. **A**) The genome of SARS-CoV-2 and the 28 predicted proteins. **B**) The workflow of proteome microarray fabrication and serum profiling on the microarray.

### Eighteen of the 28 predicted SARS-CoV-2 were prepared

To prepare recombinant proteins of SARS-CoV-2 for microarray fabrication, we first determined the amino acid sequences of predicted proteins^3^ follow a reference genome (Genbank accession No. MN908947.3). To obtain more precise analysis, we split protein S as S1 and S2^3^, and also included RBD because of its critical role during the entry of SARS-CoV-2 into the cells. The protein sequences were subjected for codon optimization and then cloned into *E*.*coli* expression vector (pET32a or pGEX-4T-1). The final expression library includes 31 clones (**Table S1**). After several rounds of optimizations, so far, we managed to purify 17 of these proteins (**Fig. S1**). As demonstrated by Western blotting with an anti-6xHis antibody and Coomassie staining, Most of the SARS-CoV-2 proteins showed clear bands of the expected size (±10 kDa) and good purity. Meanwhile, in order to cover the proteome of SARS-CoV-2 as completed as possible, and to take post-translational modification (PTM), especially glycosylation into account, we also collected recombinant SARS-CoV-2 proteins prepared using yeast cell-free system and mammalian cell expression system, from a variety of sources (**Figure S1**). Among the collected proteins, there are several different versions of S1 and N protein (**Table S1**). Finally, we obtained 37 proteins of different versions from different sources, which covers 18 out of the 28 predicted proteins of SARS-CoV-2. These proteins are suitable for microarray construction in terms of both concentration and purity.

### Protein microarray fabrication

A total of 38 proteins along with positive and negative controls were printed on the microarray slide (**Fig. 2**). Since most of the proteins were tagged with 6xHis tag, the overall quality of the microarray were evaluated by probing with an anti-6xHis antibody. The anti-6xHis antibody results showed that most of the proteins were nicely immobilized, and the microarray quality if fairly good. The detailed layout of the SARS-CoV-2 proteome microarray was indicated as well (**Fig. 2A**). High antibody responses were usually observed for COVID-19 patients while not in control sera (**Fig. 2B**). Since the Fc tag could be recognized by fluorescence labeled anti-human IgG antibody, the ACE2-Fc generated high signals for all the tests, which could serve as control for the anti-human IgG antibody, though the initial reason to include ACE2 on the microarray is for applications other than serum profiling. To test the experimental reproducibility of the serum profiling using the microarray, two COVID-19 convalescent sera were random selected. Three independent analysis for each of these two sera were repeated on the microarray. Pearson correlation coefficients between two repeats were 0.988 and 0.981 for IgG and IgM, respectively, and the overall fluorescence intensity ranges of the repeated experiments were fairly close, demonstrating high reproducibility of the microarray based serum profiling both for IgG and IgM (**Fig. 2 C-E**).

**Figure 2.**
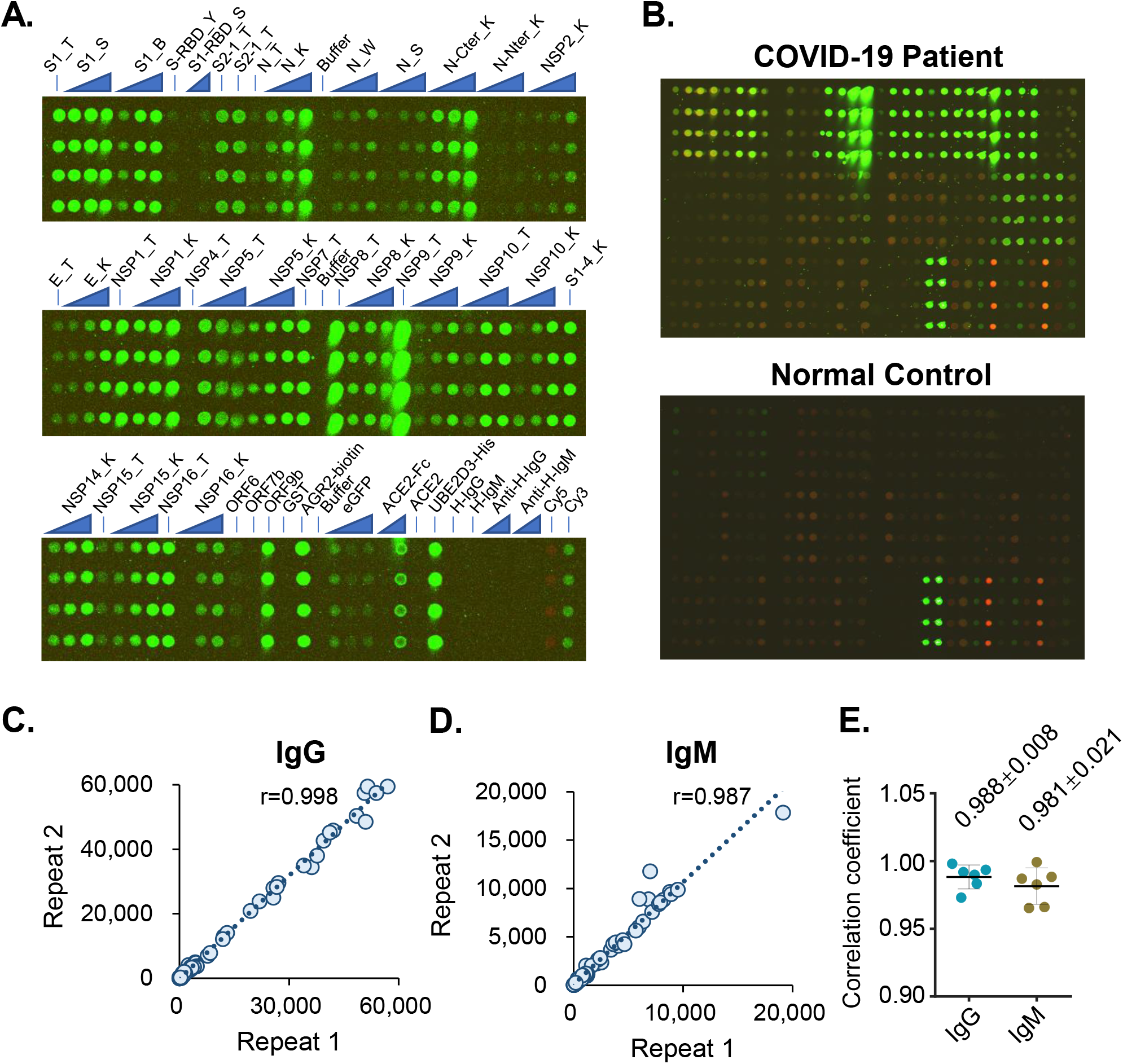
SARS-CoV-2 proteome microarray layout and quality control. **A**) There are 14 identical sub-arrays on a single microarray. A microarray was incubated with an anti-6xHis antibody to demonstrate the overall microarray quality (green). One sub-array is shown. To facilitate labeling, this sub-array is split into 3 parts. The proteins were printed in quadruplicate. The triangles indicate dilution titers of the same proteins. **B**) Representative sub-arrays probed with sera of a COVID-19 convalescent and a healthy control. The IgG and IgM responses are shown in green and red, respectively. **C**) and **D**) The correlations of the overall IgG and IgM signal intensities between two repeats probed with the same serum. **E**) Statistics of the Pearson correlation confidents among repeats probed with the same serum.

### SARS-CoV-2 specific Serum profiles depicted by proteome microarray

To globally profile the antibody response against the SARS-CoV-2 proteins in the serum of COVID-19 patients, we screened sera from 29 convalescent patients, along with 21 controls by the proteome microarray. The patients were hospitalized in Foshan Fourth hospital, China during 2020-1-25 to 2020-2-27 with variable stay time. The information of the patients was summarized in **Table 1**. Serum from each patient was collected on the day of hospital discharge when the standard criteria were met. And there is no recurrence reported for these patients. All the samples and the controls were probed on the proteome microarray, after data filtering and normalization, we built the IgG and IgM profile for each serum and performed clustering analysis to generate heatmaps for overall visualization (**Fig. 3 and Fig. 4**). The patients and controls are perfectly clustered for both IgG and IgM, justifying the utility of the SARS-CoV-2 proteome microarray for the virus specific antibody analysis of COVID-19. As expected, the N protein and S1 protein elicited high antibody responses in almost all patients but barely in control groups, confirming the efficacy of these two proteins for diagnosis. Interestingly, we also found that in some cases, some proteins other than N or S1 can generate significantly higher signals compared with that of the control groups.

**Table 1.**
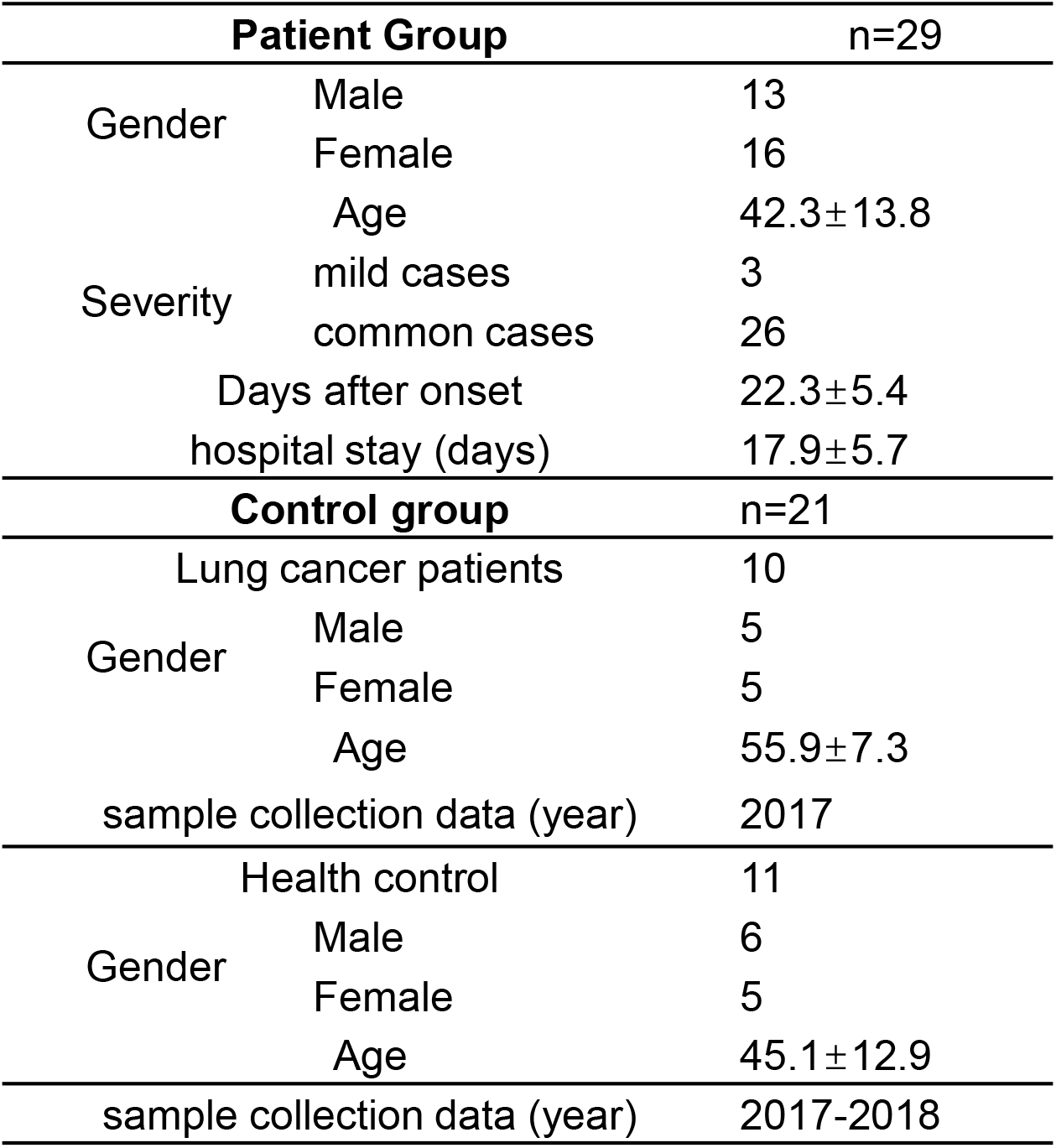
Serum samples tested in this study.

**Figure 3.**
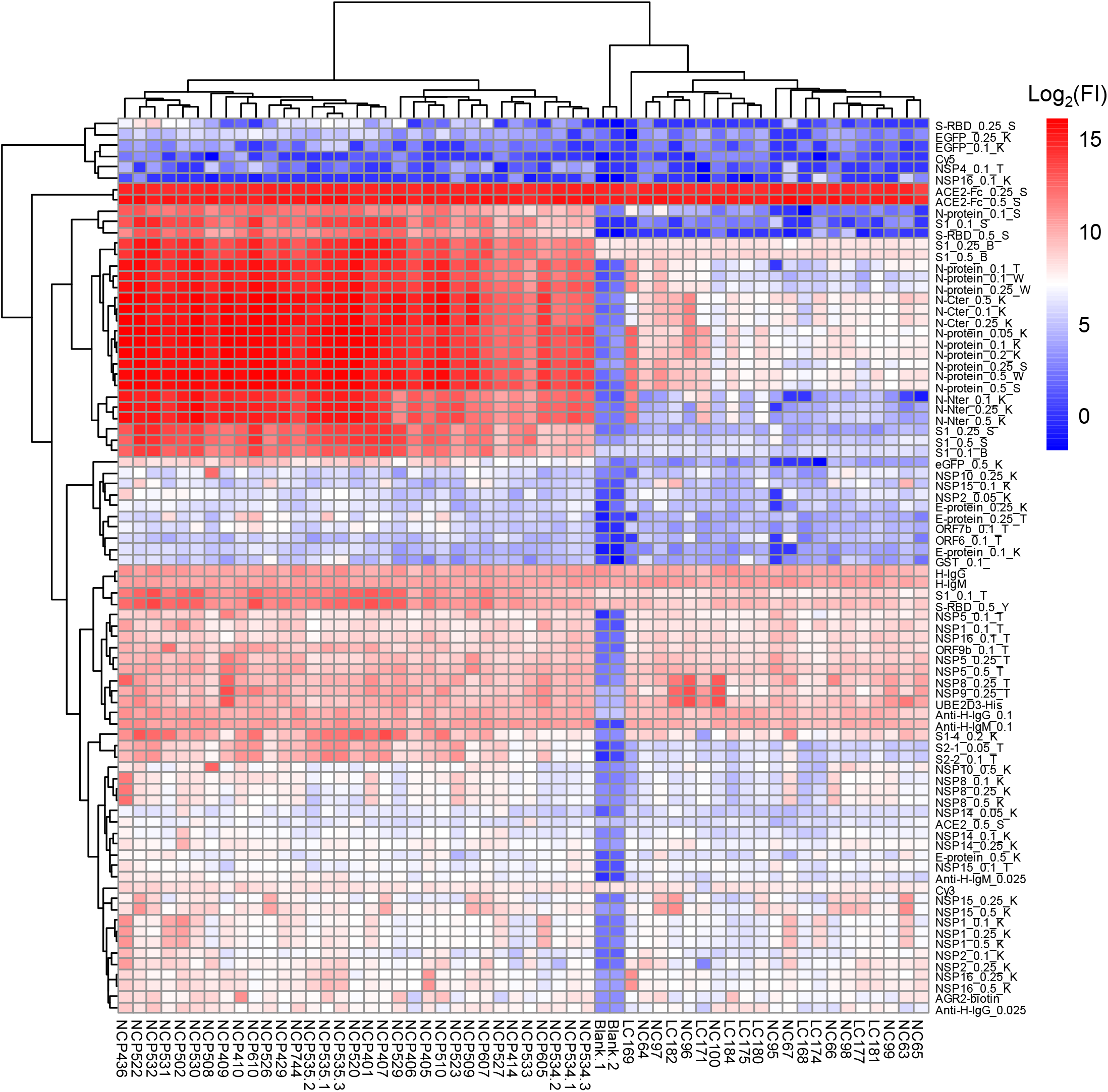
The overall SARS-CoV-2 specific IgG profiles of the 29 convalescent sera against the proteins. Each square indicates the IgG antibody response against the protein (row) in the serum (column). Proteins are shown with names along with concentrations (μg/mL) and sources. Sera are shown with group information and serum number. NCP: Novel Coronavirus Patients or COVID-19 patients; LC:Lung Cancer; NC:Normal Control. Blank means no serum. Three repeats were performed for serum NCP534 and NCP535. FI:Fluorescence Intensity.

**Figure 4.**
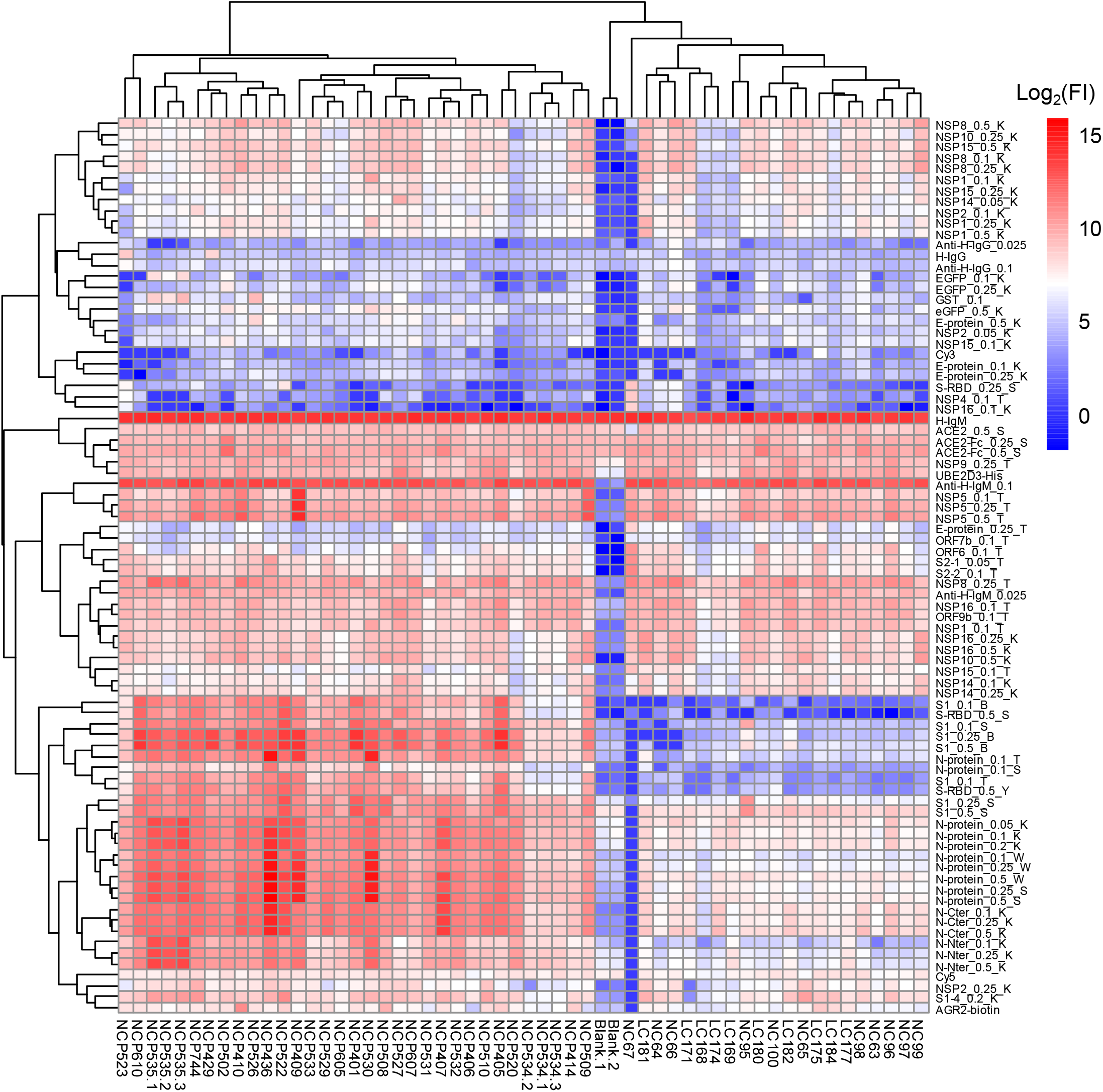
The overall SARS-CoV-2 specific IgM profiles of the 29 convalescent sera against the proteins. Each square indicates the IgM antibody response against the protein (row) in the serum (column). The rest are the same as that of **Figure 3**.

### Strong response against S and N proteins

Since S and N protein are widely used as antigens for diagnosis of COVID-19, we next characterized the serum antibody responses against these two proteins. For the present cohort, both S and N proteins, except for S1-4, were proved to have excellent discrimination ability between COVID-19 patients and controls both for IgG and IgM (**Fig. 5A, B, Fig. S3A, B**). The overall IgG signal intensities were much higher than that of IgM, mainly because the sera were collected at the convalescent stage when IgG are supposed to be dominant. It is notable that two sera from control group have significantly higher IgG antibody response to N proteins than other controls, with one to N-Nter and another to C-Nter (**Fig. 5G**), suggesting the N protein may generate a higher false positive rate than S protein, especially S1. To investigate the consistence of signal intensities among different sources or versions (C-term, N-term, domain or fragment) of proteins, we calculated the Pearson correlation coefficients among S proteins (**Fig. 5C**) and N proteins (**Fig. S2F**) using data of the convalescent sera. High correlations are observed among different concentrations of the same proteins, and the same protein from different sources is also of high correlation (**Fig. 5D, H, Fig. 2SA, B, C, G**), although N protein with high concentrations generate almost saturated signals (**Fig. S2G**). Specially, for full length S1 proteins from different sources, either obtained from *E*.*coli* (S1_T) or 293T (S1_B and S1_S) expression system, high correlations with each other were observed (**Fig. 5C, D**), indicating the proteins from different sources are all have good performance for detection. However, the background signals in control group are much lower for proteins purified from mammalian cells, *i*.*e*., 293T (**Fig. 5A**), suggesting they may possess a higher septicity and could serve as a better reagent for developing immune-diagnostics. The signals of the full length S1 protein are highly correlated with that of S-RBD (**Fig. 5E**) but with much stronger signals. In contrast, the correlation levels of S1-4 region with full-length S1 or RBD are lower (**Fig. 5C, Fig. S2D**). In addition, The S1 signals are poorly correlated with S2 proteins (**Fig. 5F**). These data may reflect the difference in immunogenicity for different regions of S protein, further epitope mapping could answer this question. Similar situations are also observed for N proteins (**Fig. 5H, Fig. S2H, I**). Interestingly, moderate but significant liner correlations were observed between IgG responses of N and S1 (**Fig. 5I**). In addition, the correlations of IgG and IgM signals for the same protein were low (**Fig. 5J, K**), this may in-part because the overall IgM signals were much lower than that of IgG (**Fig. S3**) at the convalescent stage.

**Figure 5.**
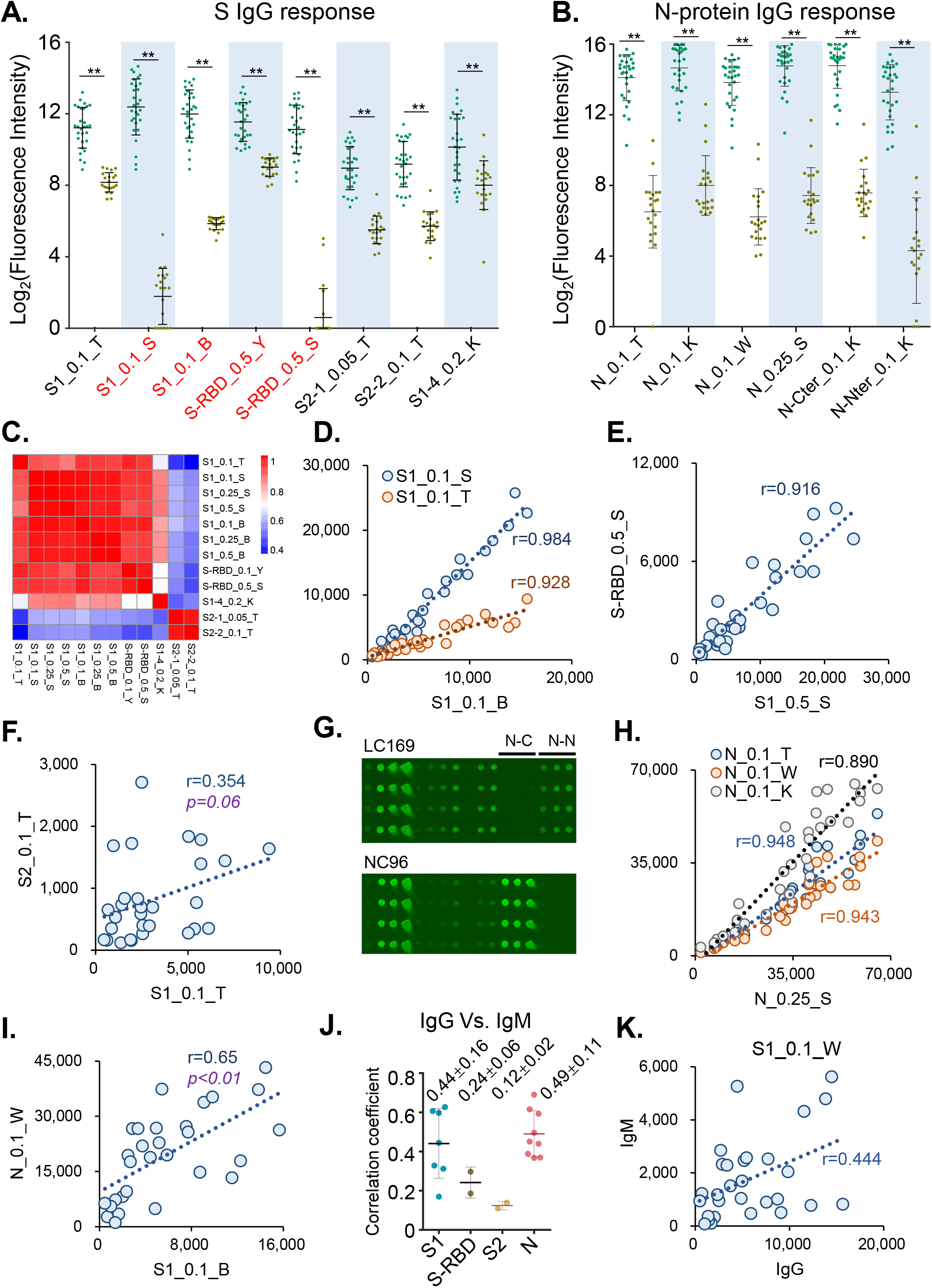
IgG response to S and N proteins. **A**) Box plots of IgG response for S1 and S2 proteins. Each dot indicates one serum sample either from convalescent group (green) or control group (brown). Mean and standard deviation value for each group are indicated. The proteins labeled with red are over expression in mammalian cell lines. P values were calculated by t.test.**, p <0.01;*, p <0.05; n.s., not significant. **B**) Box plots of IgG response for N proteins. **C**) Pearson correlation coefficient matrix of IgG response among different S1 and S2 proteins. **D-F**) Correlations of overall IgG responses among different S1 proteins (**D**), S1 vs. RBD (**E**) and S1 vs. S2 (**F**). Each spot represents one sample. **G**) One part of a sub-microarray showed the IgG responses of two controls, i.e., LC169 and NC96 against N proteins, N-Cter and N-Nter indicated the C-terminal and N-terminal of N protein, respectively. **H-I**) Correlations of the overall IgG responses among different N proteins (**H**) and N protein vs. S protein (**I**). **J**) Statistics of the Pearson correlation coefficients between IgG and IgM profile against a set of proteins. **K**) Correlations between IgG and IgM profile against S1_0.1_W.

### Antibody responses against other proteins

The proteome microarray enables us to investigate antibody responses to 18 of the 28 predicted proteins of SARS-CoV-2, including S and N proteins. As mentioned above, some proteins other than N and S proteins also generated high IgG signals (**Fig. 3**). After global analysis, we identified 6 proteins, against which high IgG responses were detected in at least one convalescent sera (**Fig. 6A**). Importantly, 44.8% (13/29) patients presented positive IgG antibody to ORF9b under the threshold set based on the signals of control sera (**Fig. 6B**). IgG antibodies to NSP5 were positive in 3/19 patients and positive in 1/21 control sera (**Fig. 6C**). To investigate if the IgG responses against ORF9b or NSP5 depends on the IgG responses against N or S protein, we calculated the correlations among them. It turned out that there are no obvious correlations between the IgG to ORF9b or NSP5 with IgG to N or S (**Fig. 6D, E**), suggesting these two proteins may provide complementary information either for diagnosis or worth further study to explore the SARS-CoV-2 specific immune response.

**Figure 6.**
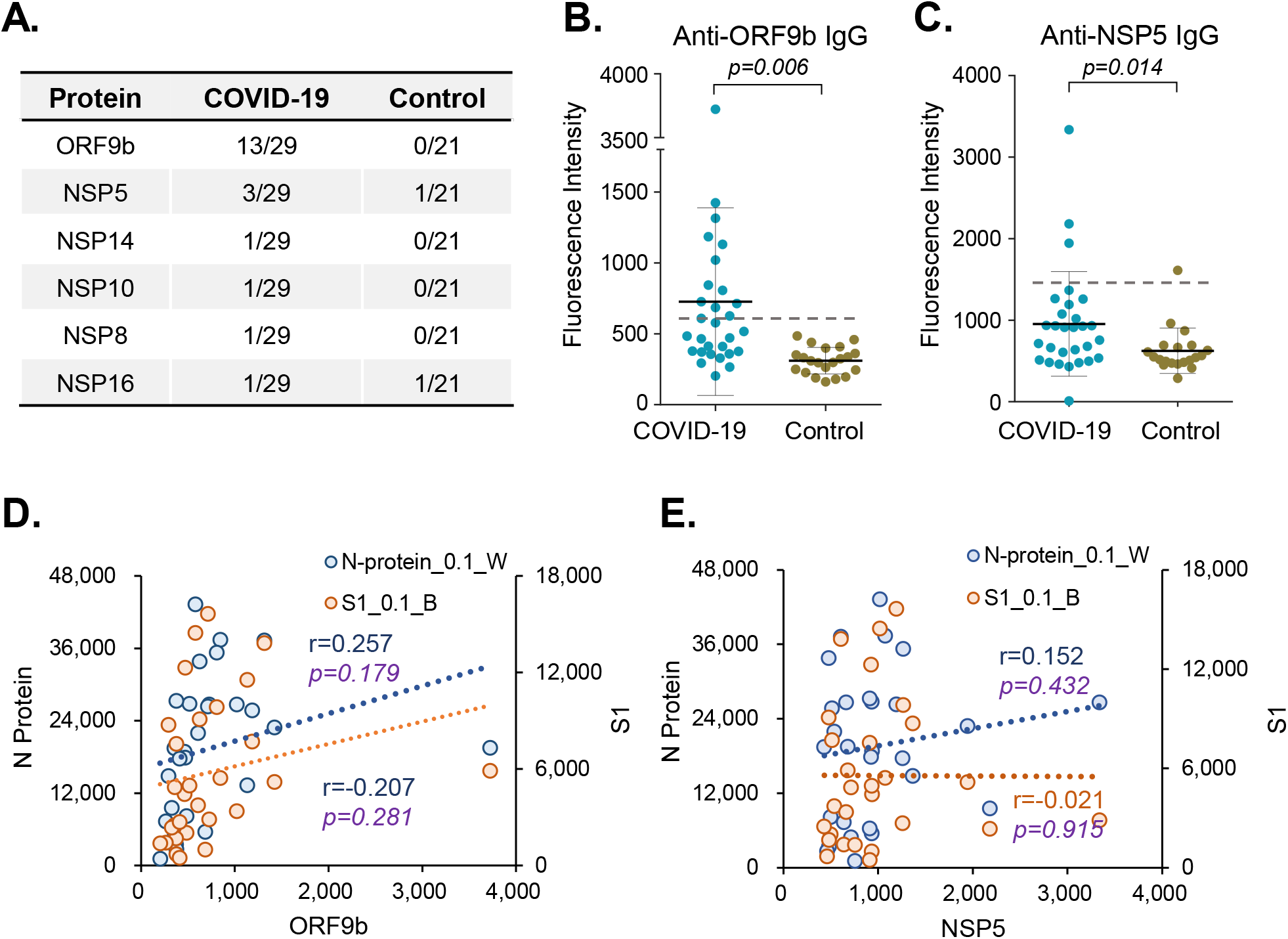
IgG response to other SARS-CoV-2 proteins. **A**) Other SARS-CoV-2 proteins that were recognized by IgG from the convalescent sera, in comparison to that of the controls. **B-C**) Anti-ORF9b IgG (**B**) or anti-NSP5 IgG (**C**) in patient and control group. Each dot represents one serum sample. Mean and standard deviation value for each group are indicated. The dashed line indicates cutoff value calculated as mean plus 3x standard deviation of the control group. P values were calculated by t test. **D-E**) Correlations of the overall IgG responses for N or S1 protein vs. ORF9b (**D**) or NSP5 (**E**).

### IgG responses are highly correlated with age, LDH and Ly%

It is known that immune response is closely related to the disease courses. To evaluate how the relationship between the antibody response and the situation of the disease, we investigated the correlations between IgG or IgM responses to proteins of virus with clinical characteristics. Not surprisingly, the days after onset were correlated with the IgG response against S1 (**Fig. 7A**) or N protein (date not shown), as the IgG response usually increases over time and reach the peak several weeks after onset, which is observed by other studies^25^ and SARS patients^26^. In contrast, there is no correlations between IgM response with days after onset. It was observed that age is also correlated with IgG response to S1 or N proteins (**Fig. 7B, Fig. S4A**). Three patients with mild symptoms indicated by red arrows show low IgG response and it is highly variable in the patients with common symptoms. It is notable that in male patients, especially older than 40, the correlation between age and S1 IgG is very poor (**Fig. 7B**). To further investigate this phenomenon, we separately analyzed the male and female patients in different age groups. For female older than 40 years old, the S1 IgG response is significantly stronger than that either in the group of female younger than 40 or the male counterpart (**Fig. 7C**). For male younger than 40, high correlation between age and S1 or N IgG response was observed (**Fig. 7D, Fig. S4C**), and this correlation is not caused by days after onset (**Fig. S4B**). For female, whatever the age range is, IgG response to S1 or N is highly correlated with age (**Fig. S4D, E**). To exclude the influence of days after onset, the patients with similar (18-24) days after onset were selected (**Fig. S4F**). High correlation is still observed (**Fig. 7E, Fig. S4G**), demonstrating the correlation is independent of days after onset.

**Figure 7.**
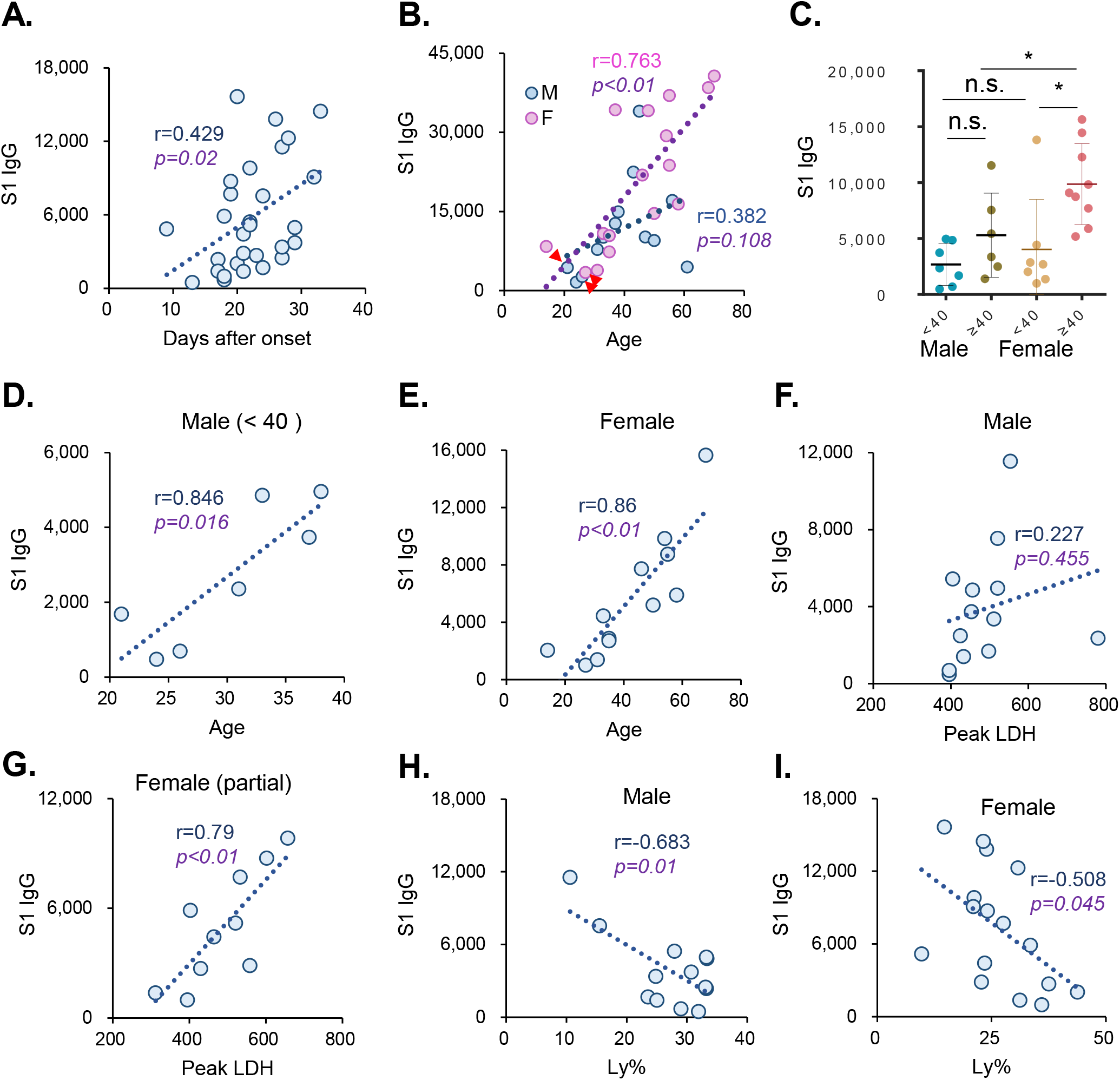
Correlation with clinical characteristics. **A**) Correlations of S1 IgG responses with Days after COVID-19 onset. Each spot indicates one COVID-19 patient. **B**)Correlations of S1 IgG responses with age either for female (blue) or male (pink). The red arrows indicate patients with mild symptoms while others with common symptoms. **C**) S1 IgG responses in groups of different age and gender. P values were calculated by t test.**, p <0.01;*, p <0.05; n.s., not significant. **D-I**) Correlations of S1 IgG responses with Age for male (age <40) (**D**), Age for female (**E**), LDH for male (**F**), LDH for female (**G**), Ly% for male (**H**) and Ly% for female (**I**). Each spot indicates one patient from the corresponding group. For **E**), only the patients with 18-24 days after onset were selected. For G), only the female patients with 18-24 days after onset and ranging in age from 20 to 60.

It was found that the IgG response against S1 or N protein is highly correlated with peak lactate dehydrogenase (LDH) level in female (**Fig. S4H**) but not significantly in male patients (**Fig. 7E**). To evaluate whether there is correlation between peak LDH and virus specific IgG response in female patients, the patients with 18-24 days after onset and ranging in age from 20 to 60 were selected to form a smaller cohort, in which, no statistical correlation between age or days after onset with peak LDH were observed (**Fig. S4I**). It turns out the correlation was still high and significant both for S1 and N protein specific IgG response **(Fig. 7G, Fig. S4J**), demonstrating the correlation is independent of age and days after onset. Since the LDH level could serve as an indicator of disease severity^26,27^, it seems that immune system of women may be more sensitive to the virus. It was also found that the IgG response to S1 or N protein negatively correlates with the percentage of lymphocyte (Ly%) (**Fig. 7H, I, Fig. S4K**), however, this correlation might be dependent of age (**Fig. S4L**).

## Materials and methods

### Construction of expression vectors

The protein sequences of SARS-CoV-2 were downloaded from GenBank (Accession number: MN908947.3). According to the optimized genetic algorithm^28^, the amino acid sequences was converted into *E*.*coli* codon-optimized gene sequences. Subsequently, the sequence optimized genes were synthesized by Sangon Biotech. (Shanghai, China). The synthesized genes were cloned into pET32a or pGEX-4T-1 and transformed into *E. coli* BL21 strain to construct the transformants. Detailed information of the clones constructed in this study is given in **Table S1**.

### Protein preparation

The recombinant proteins were expressed in E. coli BL21 by growing cells in 200 mL LB medium to an A600 of 0.6 at 37 °C. Protein expression was induced by the addition of 0.2 Mm isopropyl-β-d-thiogalactoside (IPTG) before incubating cells overnight at 16 °C. For the purification of 6xHis-tagged proteins, cell pellets were re-suspended in lysis buffer containing 50 mM Tris-HCl pH 8.0, 500 mM NaCl, 20 mM imidazole (pH 8.0), then lysed by a high-pressure cell cracker (Union-biotech, Shanghai, CHN). Cell lysates were centrifuged at 12,000 rpm for 20 mins at 4°C. Supernatants were purified with Ni2+ Sepharose beads (Senhui Microsphere Technology, Suzhou, CHN), then washed with lysis buffer and eluted with buffer containing 50 mM Tris-HCl pH 8.0, 500 mM NaCl and 300 mM imidazole pH 8.0. For the purification of GST-tagged proteins, cells were harvested and lysed by a high-pressure cell cracker in lysis buffer containing 50 mM Tris-HCl, pH 8.0, 500 mM NaCl, 1 mM DTT. After centrifugation, the supernatant was incubated with GST-Sepharose beads (Senhui Microsphere Technology, Suzhou, CHN). The target proteins were washed with lysis buffer and eluted with 50 mM Tris-HCl, pH 8.0, 500 mM NaCl, 1 mM DTT, 40 mM glutathione. The purified proteins were analyzed by SDS-PAGE followed by Western blotting using an anti-His antibody (Merck millipore, USA) and Coomassie brilliant blue staining. Recombinant SARS-CoV-2 proteins were also collected from commercial sources. Detailed information of the recombinant proteins prepared in this study is given in **Table S1**.

### Protein microarray fabrication

The proteins, along with negative (BSA) and positive controls (anti-Human IgG and IgM antibody), were printed in quadruplicate on PATH substrate slide (Grace Bio-Labs, Oregon, USA) to generate identical arrays in a 2 × 7 subarray format using Super Marathon printer (Arrayjet, UK). Protein arrays were stored at -80°C until use.

### Patients and samples

The Institutional Ethics Review Committee of Foshan Fourth Hospital, Foshan, China approved this study, and written informed consent was obtained from each patient. COVID-19 patients were hospitalized and received treatment in Foshan Forth hospital during 2020-1-25 to 2020-2-27 with variable stay time (**Table 1**). Serum samples were collected when the patients were discharged from the hospital. Sera of control group from Lung cancer patients and healthy controls were collected from Ruijin Hospital, Shanghai, China. All sera were stored at -80 °C until use.

### Microarray based serum analysis

A 14-chamber rubber gasket was mounted onto each slide to create individual chambers for the 14 identical subarrays. The microarray was used for serum profiling as described previously^29^ with minor modifications. Briefly, the arrays stored at -80°C were warmed to room temperature and then incubated in blocking buffer (3% BSA in PBS buffer with 0.1% Tween 20) for 3 h. Serum samples were diluted 1:200 in PBS containing 0.1% Tween 20. A total of 200 μL of diluted serum or buffer only was incubated with each subarray overnight at 4°C. The arrays were washed with PBST and bound autoantibodies were detected by incubating with Cy3-conjugated goat anti-human IgG and Alexa Fluor 647-conjugated donkey anti-human IgM (Jackson ImmunoResearch, PA, USA), the antibodies were diluted 1: 1,000 in PBST, and incubated at room temperature for 1 h. The microarrays were then washed with PBST and dried by centrifugation at room temperature and scanned by LuxScan 10K-A (CapitalBio Corporation, Beijing, China) with the parameters set as 95% laser power/ PMT 550 and 95% laser power/ PMT 480 for IgM and IgG, respectively. The fluorescent intensity data was extracted by GenePix Pro 6.0 software (Molecular Devices, CA, USA).

### Statistics

Signal Intensity was defined as median of foreground subtracted by median of background for each spot and then averaged of the quadruplicate spots for each protein. IgG and IgM data were analyzed separately. Before processing, data from some spots, such as NSP7_0.1_T, NSP9P_K, are excluded for probably printing contamination. Pearson correlation coefficient between two proteins or indicators and the corresponding *p* value was calculated by SPSS software under the default parameters. Cluster analysis was performed by pheatmap package in R^30^.

## Discussion

In order to profile the SARS-CoV-2 specific IgG/IgM responses, we have constructed a SARS-CoV-2 proteome microarray with 18 of the 28 predicted proteins. To our knowledge, this is the first of such. A set of 29 recovered sera were analyzed on the microarray, global IgG and IgM profile were obtained simultaneously through a dual color strategy. Our data clearly showed that both protein N and S1 are suitable for diagnostics, while S1 purified from mammalian cell may possess better specificity. Significant antibody responses were identified for ORF9b and NSP5. We showed that the level of S1 IgG positively correlate to age and the level of LDH while negatively correlate to Ly%.

The SARS-CoV-2 proteome microarray enables not only the global profiling of virus specific antibody responses but also providing semi-quantitative information. By adopting the dual color strategy of microarray, we can measure IgG and IgM simultaneously. For the convalescent COVID-19 patients tested in this study that with a median of 22 days after onset, we found that the overall IgG response is significantly higher than that of IgM, indicating for these patients the SARS-CoV-2 specific IgG responses are dominant at the convalescent phase, although IgM level might reach the peak at a similar time point with that of IgG, according to some studies of SARS-CoV^31,32^.

It is well known that S1 and N proteins are the dominant antigens of SARS-CoV and SARS-CoV-2 that elicit both IgG and IgM antibodies, and antibody response against N protein is usually stronger. However, we found for two of the control sera, strong IgG bindings were observed for N protein, and specifically one control recognizing N protein at the N terminal while another at the C terminal. This maybe due to the high conservation of N protein sequences across the coronavirus species, this indicating we should be aware of the false positive when applying N protein for diagnosis. In contrast, S1 protein demonstrating a higher specificity. Thus an ideal choice of developing immune-diagnostics maybe the combining of both N protein and S1 protein.

We also compared the antibody responses against a variety versions of S1, including the full length, the RBD domain, the N terminal and the C terminal. The antibody response to RBD region is highly correlated with that to full length protein but with weaker signals, however, the correlations among other S1 versions are not significant, suggesting dominant epitopes that elicit antibodies might differ among individuals. Further study of detailed epitope mapping might give us a clear answer.

In this study, we also found the significant presence of IgG and IgM against ORF9b (13 out of 29 cases) and NSP5 (3 out of 29 cases). ORF9b is predicted as an accessory protein, exhibiting high overall sequence similarity to SARS and SARS-like COVs ORF9b (V23I)^3^, and is likely to be a lipid binding protein^33^. Previous studies showed that SARS ORF9b suppresses innate immunity by targeting mitochondria^34^. Two previous studies have found antibody against SARS ORF9b presented in the sera of patients recovering from SARS^35,36^. Our study also demonstrates the potential of antibody against ORF9b for detection of convalescent COVID-19 patients. COVID-19 NSP5 is also highly homologous to SARS NSP5 (96% identity, 98% similarity). Its homologous proteins in a variety of coronaviruses have been proven to impair IFN response^37-39^. Our study is the first report to provide experimental evidence to show the existence of NSP5 specific antibody in convalescents. Since NSP5 is a non-structural protein, theoretically, it should present only in the infected cells but not in virions. So antibody against NSP5 has the potential to be applied to distinguish between COVID-19 patients and healthy people immunized with inactivated virus.

We have analyzed the correlations between the COVID-19 specific IgG responses with clinical characteristics as well. It is expected that IgG responses improve over time within one or two months after onset^25,31,32^ and we did observe a significant correlation between IgG signals with days after onset. We also found peak LDH was highly correlated with IgG response, especially for female patients. As many studies reported, LDH tends to have a higher level in severe COVID-19 patients and could be an indicator of severity^26,27^. In fact, it has been observed in SARS patients that more severe SARS is associated with more robust serological response^26,40^, similar association was confirmed in COVID-19 patients, especially for females. Interestingly, we observed high correlation between age with IgG response in female patients and in male patients with age less than 40 but not in older male patients, implying the humoral response against SARS-CoV-2 may differ in gender. It is reported that severe cases are significantly more frequent in aged patients and the mortality of male patients is higher than that of female, but the reason is not clear. Based on our observations, we assumed that the situation might be associated with the immune response. However, female patients, compared with male patients, may generate humoral response more efficiently. This difference should be considered during treatment.

There are some limitations of the current SARS-CoV-2 proteome microarray. Firstly, due to the difficulty of protein expression and purification, there are still 10 proteins missing^3^. We will try to obtain these proteins through vigorous optimization or other sources, interesting finding is anticipated in the near future for these missing proteins. Secondly, most of the proteins on the microarray are not expressed in mammalian cells, critical post-translational modifications, such as glycosylation is absent. It is known that there are 23 N-glycosylation sites on S protein, which is heavily glycosylated, and the glycosylation may play critical roles in antibody-antigen recognition^5,41^. We are preparing these proteins using mammalian cell systems. Once the microarray is upgraded with proteins purified from mammalian cells, PTM specific IgG/IgM response may could be elicited. Thirdly, only 29 samples at collected at a single time point were analyzed. Though there are some interesting findings, we believe some of the current conclusions could be strengthened by including more samples. Furthermore, longitudinal samples^29,42^ collected at different time points from the same individual after diagnosis or even after cured may enable us to reveal the dynamics of the SARS-CoV-2 specific IgG/IgM responses. The data may could be further linked to the severity of COVID-19 among different patients.

The application of the SARS-CoV-2 proteome microarray is not limited to serum profiling. It could also be explored for host-pathogen interaction^43^, drug/ small molecule target identification^44,45^,and antibody specificity assessment^46^.

Through the same construction procedure, we could easily expand the microarray to a pan human coronavirus proteome microarray by including the other two severe coronaviruses, *i*.*e*., SARS-CoV^11,47,48^ and MERS-CoV^47^, as well as the four known mild human coronaviruses^49,50^, *i*.*e*., CoV 229E, CoV OC43, CoV HKU-1 and CoV NL63. By applying this microarray, we can assess the immune response to coronavirus on a systems level, and the possible cross-reactivity could be easily judged.

Taken together, we have constructed the first SARS-CoV-2 proteome microarray, this microarray could be applied for a variety of applications, including but not limited to in-depth IgG/ IgM response profiling. Through the analysis of convalescent sera on the microarray, we obtained the first overall picture of SARS-CoV-2 specific IgG/ IgM profile. We believe that the findings in this study will shed light in the development of more precise diagnostic kit, more appropriate treatment and effective vaccine for combating the global crisis that we are facing now.

### Funding sources

This work was partially supported by National Key Research and Development Program of China Grant (No. 2016YFA0500600), National Natural Science Foundation of China (No. 31970130, 31600672, 31670831, and 31370813).

## Data Availability

The SARS-CoV-2 proteome microarray data are deposited on Protein Microarray Database (http://www.proteinmicroarray.cn) under the accession number PMDE241. Additional data related to this paper may be requested from the authors.

http://www.proteinmicroarray.cn/index.php/experiment/detail?experiment_id=241

## Acknowledgements

We thank Dr. Min Guo of Healthcode Co., Ltd. for providing affinity purified proteins. We thank Dr. Guo-Jun Lang of Sanyou Biopharmaceuticals Co., Ltd. for provide proteins and antibodies. We also thank Dr. Jie Wang of VACURE l Biotechnology Co.,Ltd., Dr. Yin-Lai Li of Hangzhou Bioeast biotech. Co.,Ltd., and Sino biological Co.,Ltd. for providing the proteins.

## Author contributions

S-C. T. developed the conceptual ideas and designed the study. J. Z., W. W., D. M. and X. Y. collected the sera samples and provided key reagents. H-W. J., Y. L, H-N. Z., H. Q. performed the experiments, S-C.T., Y. L., and H-W. J. wrote the manuscript with suggestions from other authors.

---

**Figure S1.**
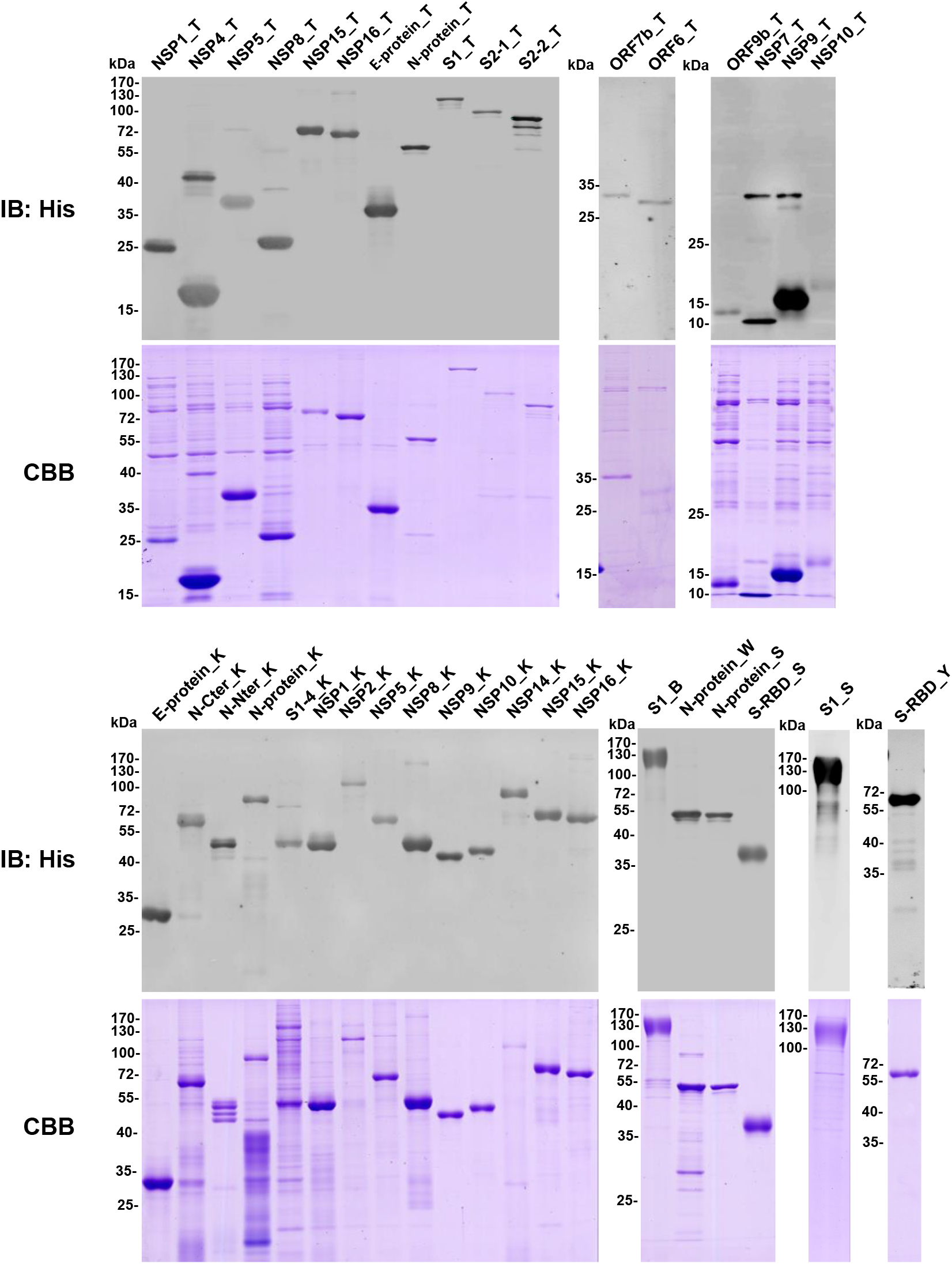
The SARS-CoV-2 proteins included in this proteome microarray. The Up panel is western blotting with an anti-6xHis antibody. The low panel is Coomassie staining. These proteins were prepared and collected from different sources. **T**: Tao Lab (our laboratory); **B**: Hangzhou Bioeast biotech. Co., Ltd.; **K**: Healthcode Co., Ltd.; **S**: Sanyou biopharmaceuticals Co., Ltd.; **W**: VACURE l Biotechnology Co., Ltd. **Y**: Sino biological Co., Ltd.

**Figure S2.**
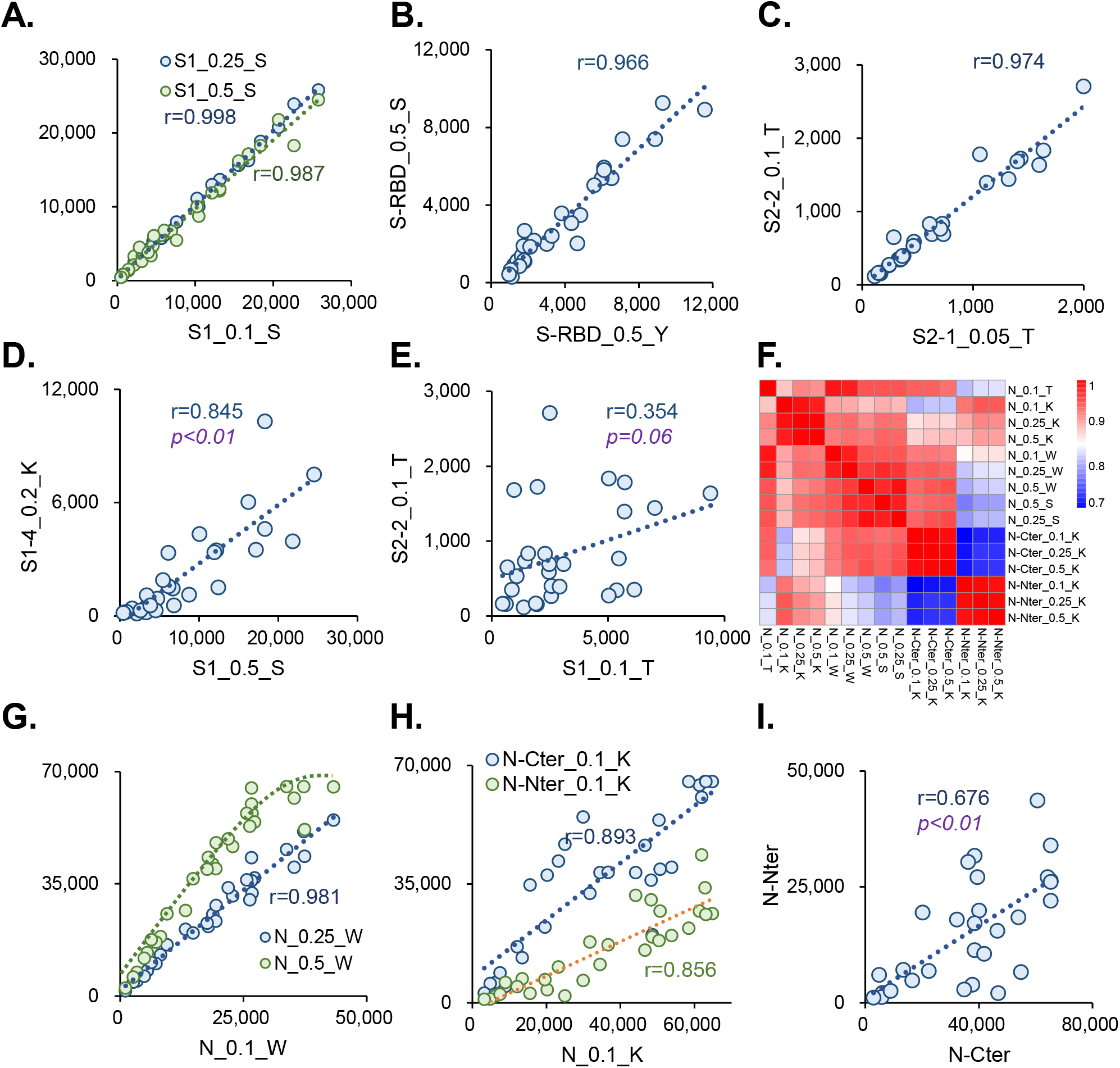
IgG response to S and N proteins. **A-E**)Correlations of the overall IgG responses among different dilutions of protein S1_S (**A**), two RDB proteins from different sources (**B**), S2-1 vs. S2-2 (**C**), S1 vs. S1-4 (**D**) and S1 vs. S2 (**E**). **F**) Pearson correlation coefficient matrix of IgG responses among N proteins. Each spot represents one sample. **G-I**) Correlations of the overall IgG responses among different dilutions of protein N_W (**A**), N-Nter/ N-Cter vs. N protein (**H**) and N-Cter vs. N-Nter (**I**).

**Figure S3.**
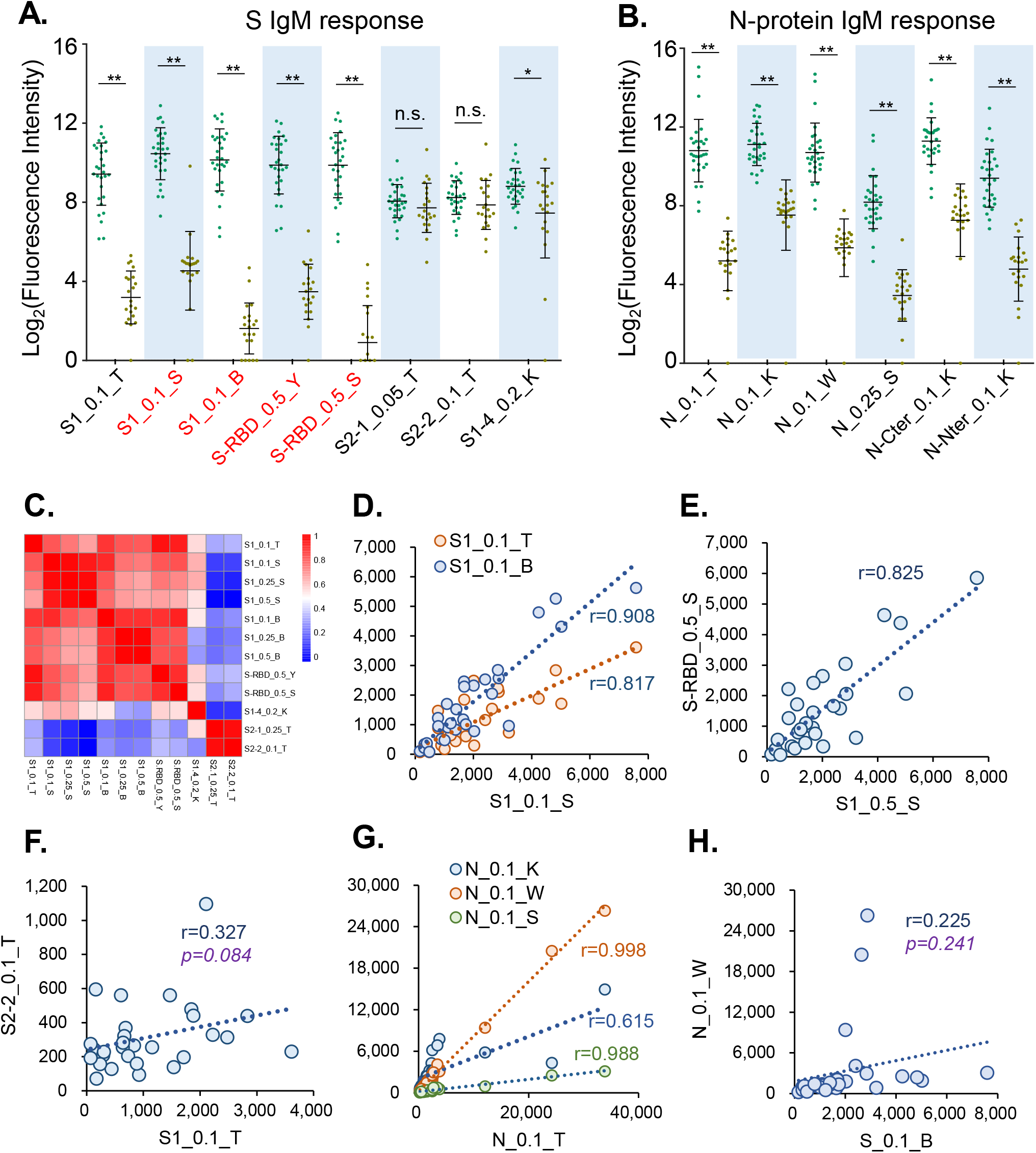
IgM Antibody response to S and N proteins. **A**) Box plots of IgM responses to S1 and S2 proteins. Each dot indicates one serum sample either from patient group (green) or control group (brown). Mean and standard deviation for each group are indicated. The proteins labeled with red are over-expressed in 293T. P values were calculated by t test.**, p <0.01;*, p <0.05; n.s., not significant. **B**) Box plots of IgM responses to N proteins. **C**) Pearson correlation coefficient matrix of IgM responses among S1 and S2 proteins of different versions from different sources. (**D-H**) Correlations of the overall IgM responses among S1 proteins (**D**), S1 vs. RBD (**E**), S1 vs. S2 proteins (**F**), different N proteins (**G**) and N vs. S proteins (**H**). Each spot represents one sample.

**Figure S4.**
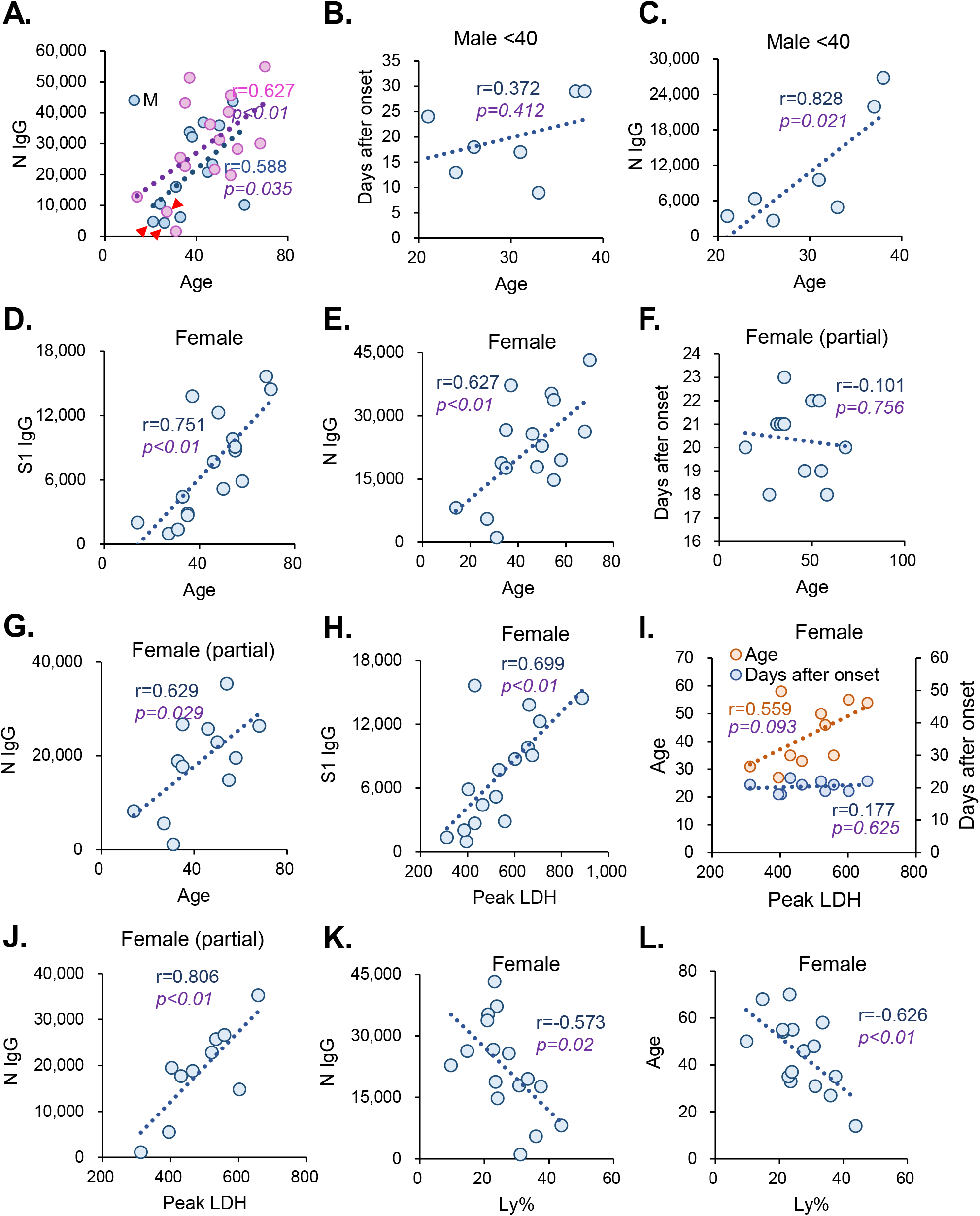
Correlation with clinical characteristics. **A**) Correlations of N protein specific IgG responses with days after onset. The red arrows indicate patients with mild symptoms while others with regular symptoms. **B-L**) Correlations of S1 specific IgG or N specific IgG with Days after onset, Age, LDH or Ly% in different groups, specifically, Days after onset vs. Age for male (age <40) (**B**), N protein specific IgG vs. Age for male (age <40)(C), S1 protein specific IgG vs. Age for female (**D**), N protein specific IgG vs. Age for female (**E**), Days after onset vs. Age for female patients with18-24 days after onset (**F**), N protein specific IgG vs. Age for female patients with18-24 days after onset (**G**), S1 protein specific IgG vs. LDH for female; (**H**), Age or Days after onset vs. LDH for female patients with 18-24 days after onset and ranging in age from 20 to 60. (**I**), N protein specific IgG vs. LDH for female patients with 18-24 days after onset and ranging in age from 20 to 60. (**J**), N protein specific IgG vs. Ly% for female (**K**) and Age vs. Ly% for female (**L**). For **F**) and **G**), only the patients with 18-24 days after onset were selected.

